# Multiple synchronous spinal dural arteriovenous fistulas: A systematic literature review

**DOI:** 10.1101/2023.08.21.23294395

**Authors:** Yusuke Ikeuchi, Atsushi Fujita, Masaaki Kohta, Shunsuke Yamanishi, Kazuhiro Tanaka, Takashi Sasayama

## Abstract

**Background:** Multiple spinal dural arteriovenous fistulas (SDAVFs) are rare and account for only 1–2% of all SDAVF cases. The treatment for SDAVFs typically involves either direct surgy or endovascular treatment. Identifying the precise location of all fistulas is paramount for successful treatment procedures when multiple SDAVFs are present. They can be classified as synchronous (occurring simultaneously) or metachronous (occurring at different times), with each type differing with respect to etiology, diagnosis, and treatment. This study systematically reviewed the literature on multiple synchronous SDAVFs.

**Methods:** A comprehensive search was performed to identify all published multiple synchronous SDAVF cases. Overall, 23 patients with multiple SDAVFs were identified, including 21 from 19 articles and 2 from this study. Clinical presentation, lesion location, radiographic features, surgical treatment, and outcome were analyzed in each patient.

**Results:** All patients in this study were male individuals, and the duration from symptom onset to diagnosis in many of these patients was longer than that previously reported. Previous studies suggested that multiple SDAVFs typically occurred within three of fewer vertebral levels. However, >50% of examined patients had remote lesions separated by more than three vertebral levels. Patients with remote lesions had a significantly worse outcome (1/7 vs. 8/11, P=0.049). Therefore, accurate localization of fistulas before spinal angiography is critical for managing multiple remote SDAVFs.

**Conclusions:** Considering the possibility of multiple remote SDAVFs, careful interpretation of imaging findings is essential for accurate diagnosis and appropriate treatment planning.

**KEY MESSAGES:** **What is already known on this topic**

Multiple spinal dural arteriovenous fistulas (SDAVFs) can be classified as synchronous (occurring simultaneously) or metachronous (occurring at different times), and they differ in etiology, diagnosis, and treatment. However, the differences between two conditions need to be better clarified.

**What this study adds**

Patients with remote SDAVFs tended to receive more treatments and had worse prognosis than those with non-remote SDAVFs. Therefore, correct localization of fistulas before spinal angiography is essential for managing multiple remote SDAVFs.

**How this study might affect research, practice or policy**

Multiple remote SDAVFs may occur in patients; therefore, careful interpretation of imaging results is critical for accurate diagnosis and appropriate treatment planning.

## INTRODUCTION

Spinal dural arteriovenous fistulas (SDAVFs) are acquired pathological arteriovenous shunts that develop in the layers of the dura and account for approximately 80% of all spinal vascular malformations.^1^ They often affect the spinal cord venous drainage, resulting in a slowly progressive neurological deterioration with paraparesis, bowel and bladder dysfunction, and sensory disturbances.^2^ The standard treatment methods for SDAVFs usually involve either direct surgical procedures or endovascular therapies. Before treatment, spinal angiography is typically conducted to pinpoint the shunt location and analyze the vasculature, which aids in devising appropriate treatment plans. Traditionally, comprehensive spinal angiography is conducted; however, this approach is time consuming, places a significant burden on patients, and carries high examination risks. Consequently, recent practices have often employed computed tomography angiography (CTA) or magnetic resonance angiography (MRA) to identify the shunt site, restricting spinal angiography to the immediate vicinity of the shunt.

Furthermore, some case studies have reported only a few patients with multiple SDAVFs.^3–5^ Two types of multiple SDAVFs have been defined, namely synchronous (two or more SDAVFs occurring simultaneously) and metachronous (SDAVFs occurring at different times). These differ in etiology, diagnostic procedures, and treatment.^6^ However, both types of multiple SDAVFs need to be clarified.

The present study aimed to review the literature on multiple synchronous SDAVFs according to the Preferred Reporting Items for Systematic Reviews and Meta-Analyses (PRISMA) guidelines and present two cases from our institution.

## METHODS

### Literature review

PubMed, Web of Science, and Cochrane databases were searched using the following terms: “spinal dural arteriovenous fistula” or “spinal dural arterio-venous fistula” in combination with “multiple,” “double,” “triple,” “concurrent,” “synchronous,” “concomitant,” “simultaneous” or “bilateral.” We did not limit our search to a specific timeframe and included only articles written in English. This study was conducted in accordance with PRISMA guidelines, and the collected articles were integrated into an EndNote library. Editorials, letters to the editor, and commentaries were excluded; 338 articles were initially identified. The article review phases involved initial title screening, abstract screening, and full-text review. Two independent reviewers performed the full-text reviews. The inclusion criteria were as follows: 1) studies including patients who underwent operative management or neuroendovascular therapy of multiple synchronous SDAVFs and 2) articles with sufficient information regarding the site of lesions and outcomes for each patient to be disaggregated. The exclusion criteria were as follows: 1) studies on double DAVFs, which were intracranial and spinal lesions; 2) studies on craniocervical DAVF; and 3) studies on epidural or perimedullary AVF or arteriovenous malformation. All relevant articles on multiple SDAVFs were reviewed. No additional patients were identified in the references of the included studies.

Twenty-one patients with multiple SDAVFs were identified in 19 studies.^1,3,5,7–22^ With the inclusion of the 2 cases in the present study, 23 patients were identified in total (Figure 1). Factors such as clinical presentation, lesion location, radiographic features, surgical treatment, and outcome were analyzed in each patient (Table 1).

**Figure 1.**
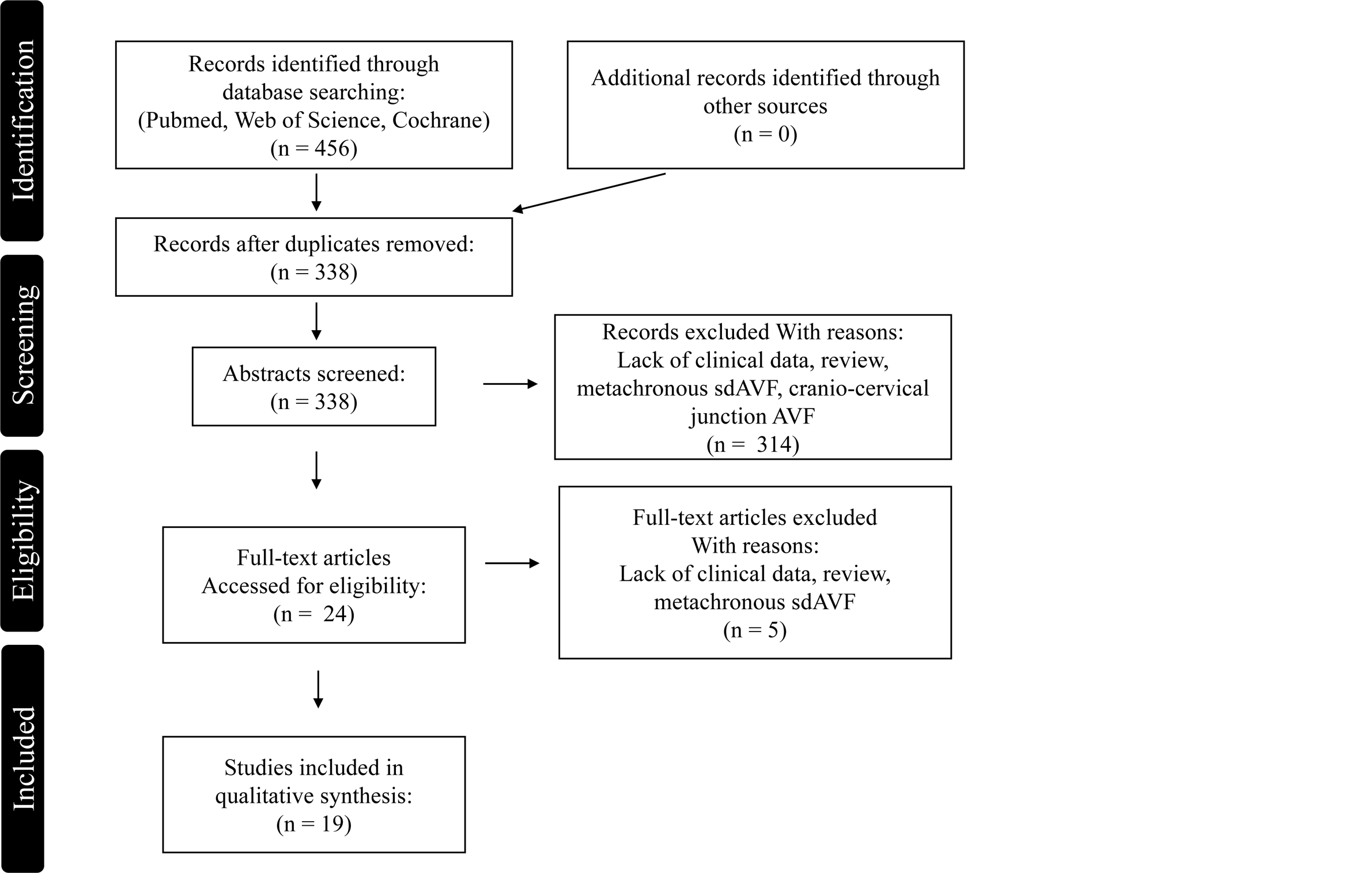
PRISMA flow diagram of article selection for meta-analysis. AVF, arteriovenous fistula; SDAVF, spinal dural arteriovenous fistula.

**Table 1.**
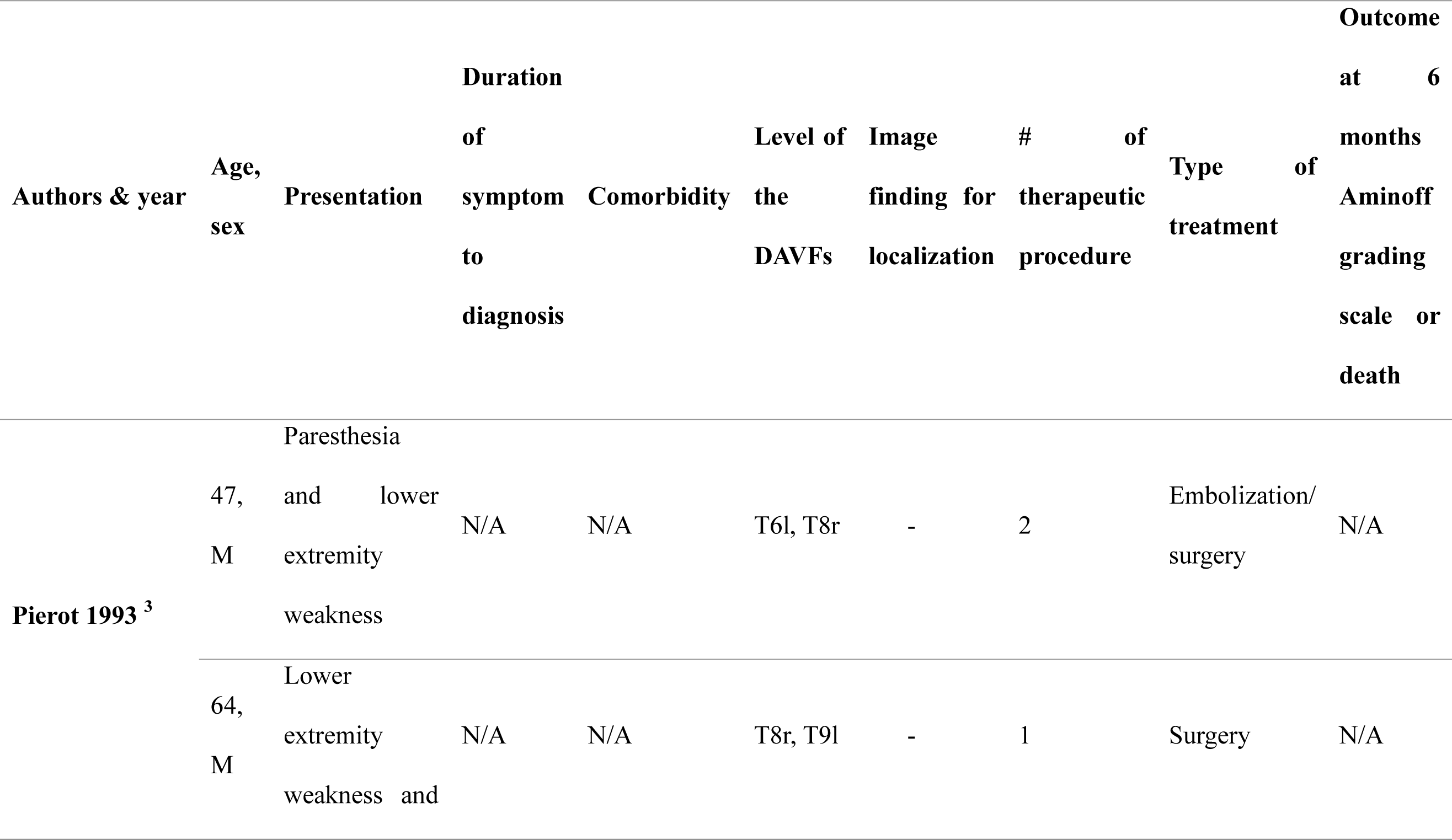
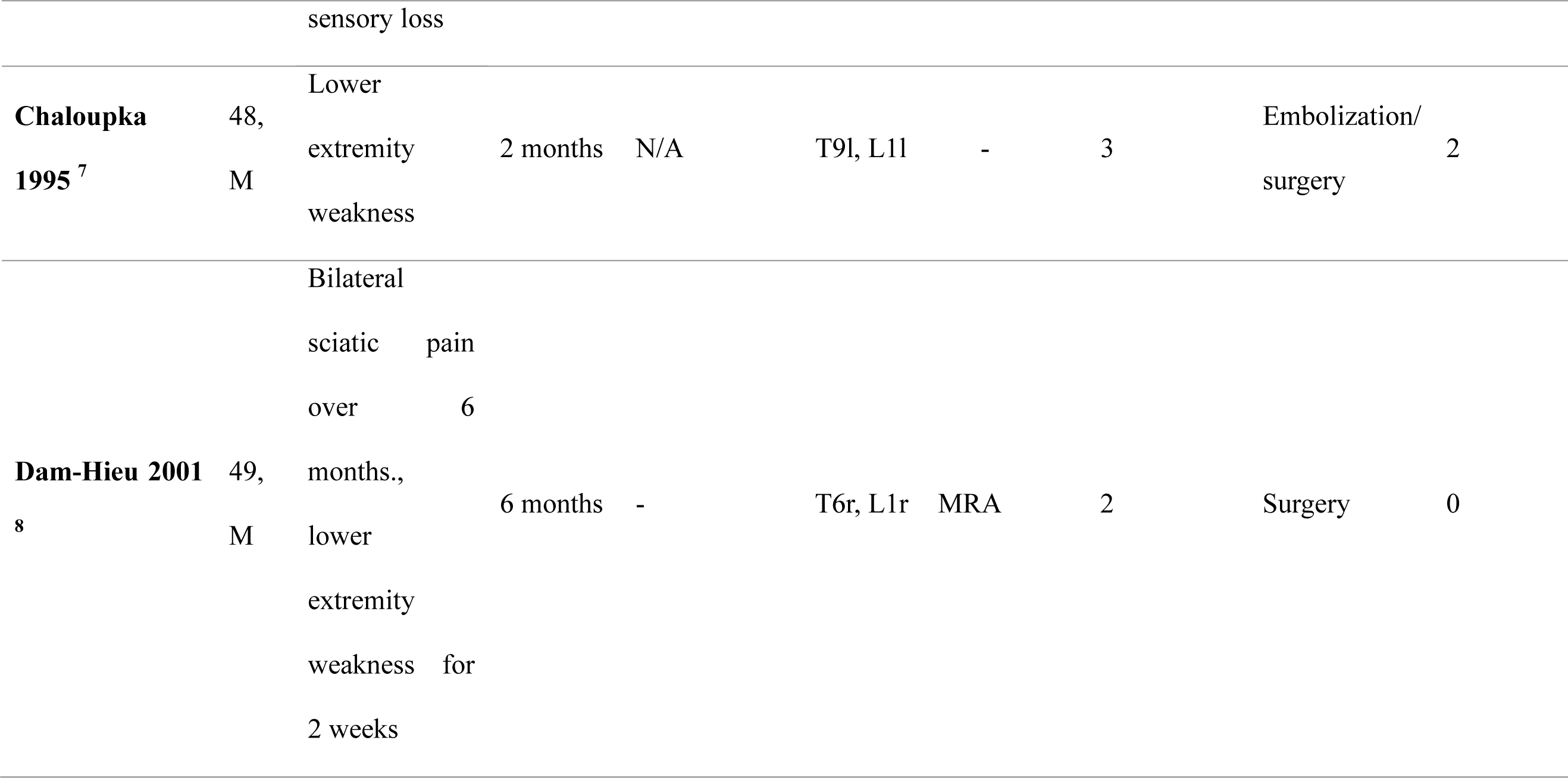
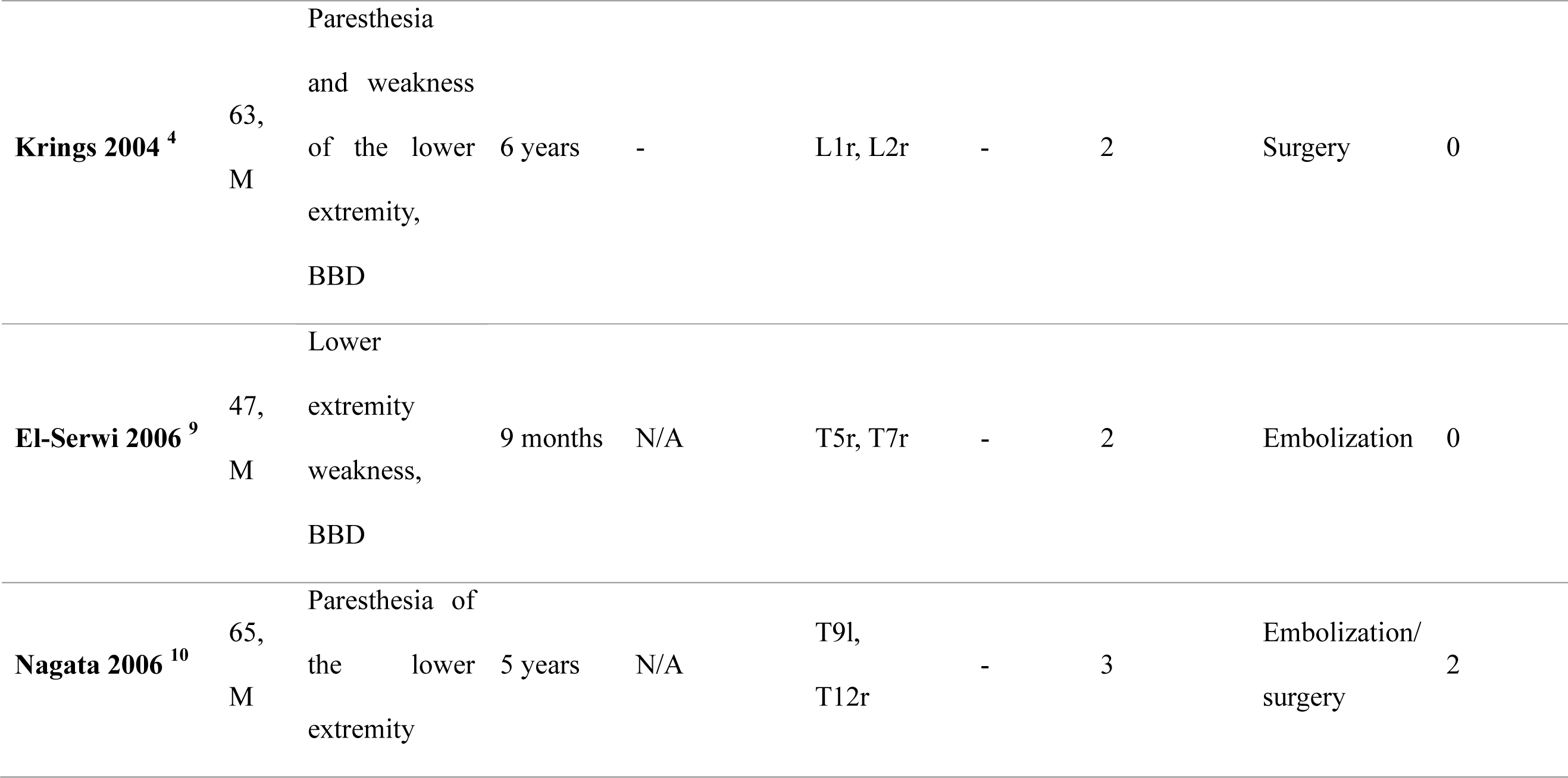
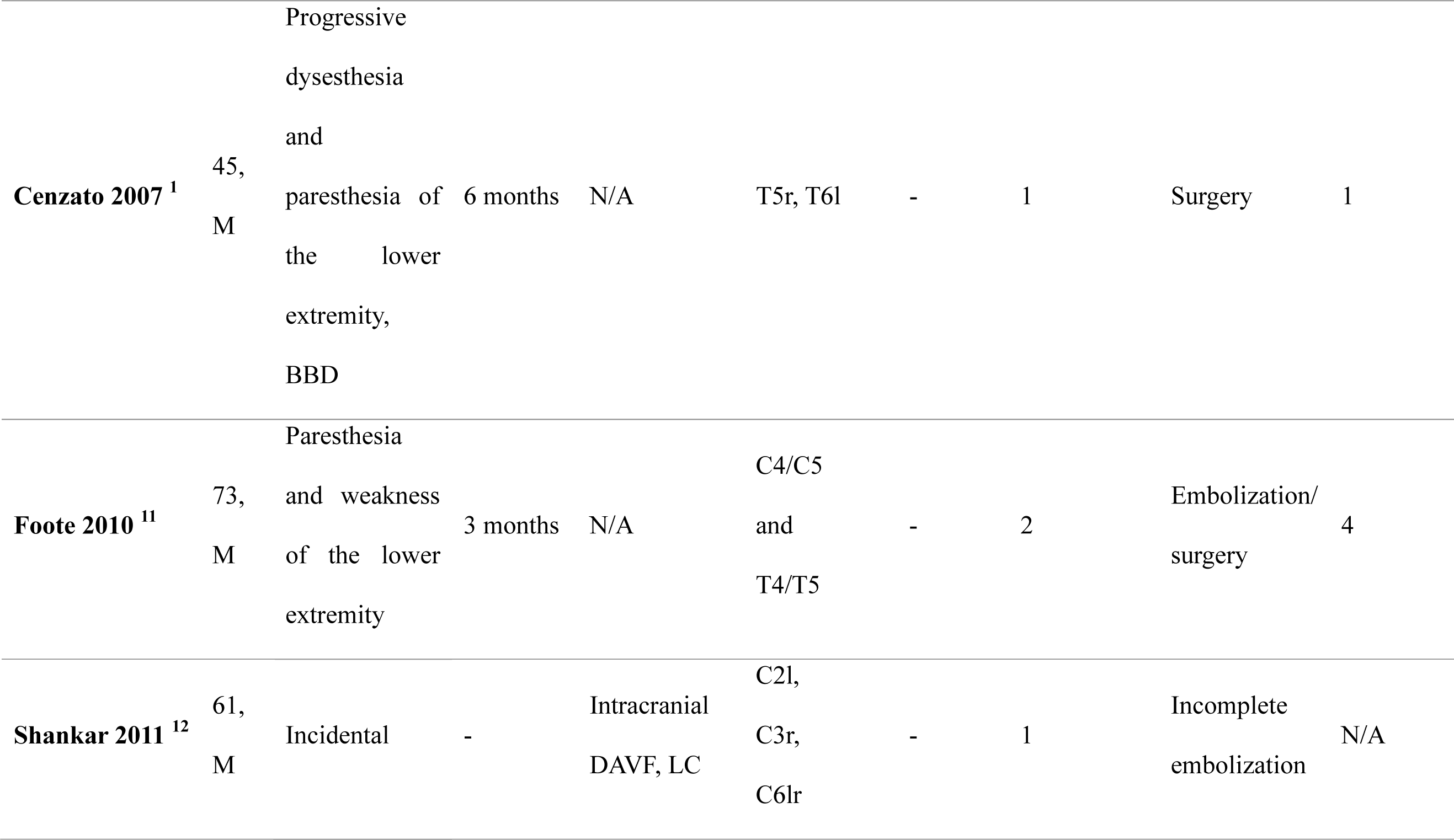
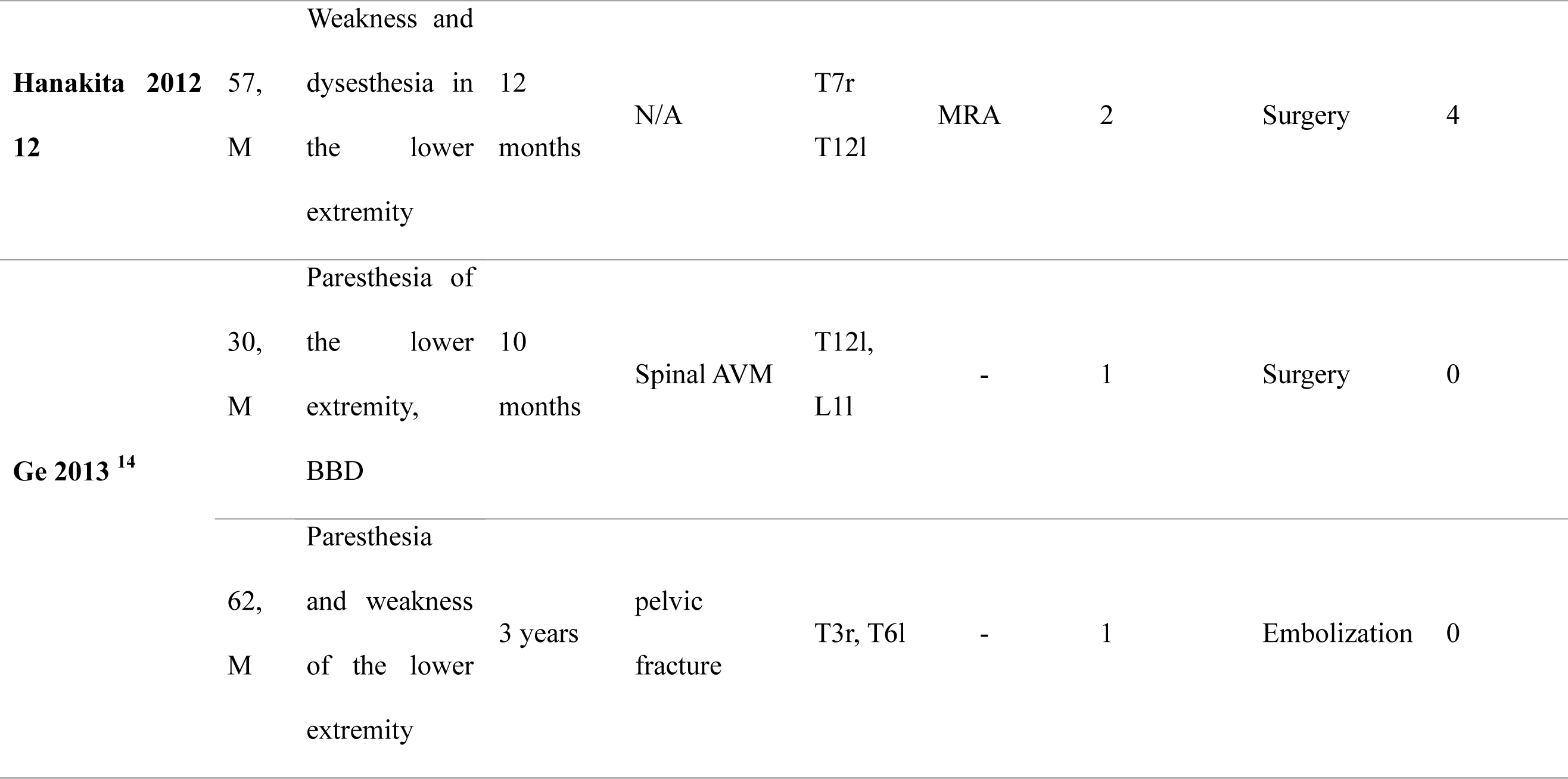
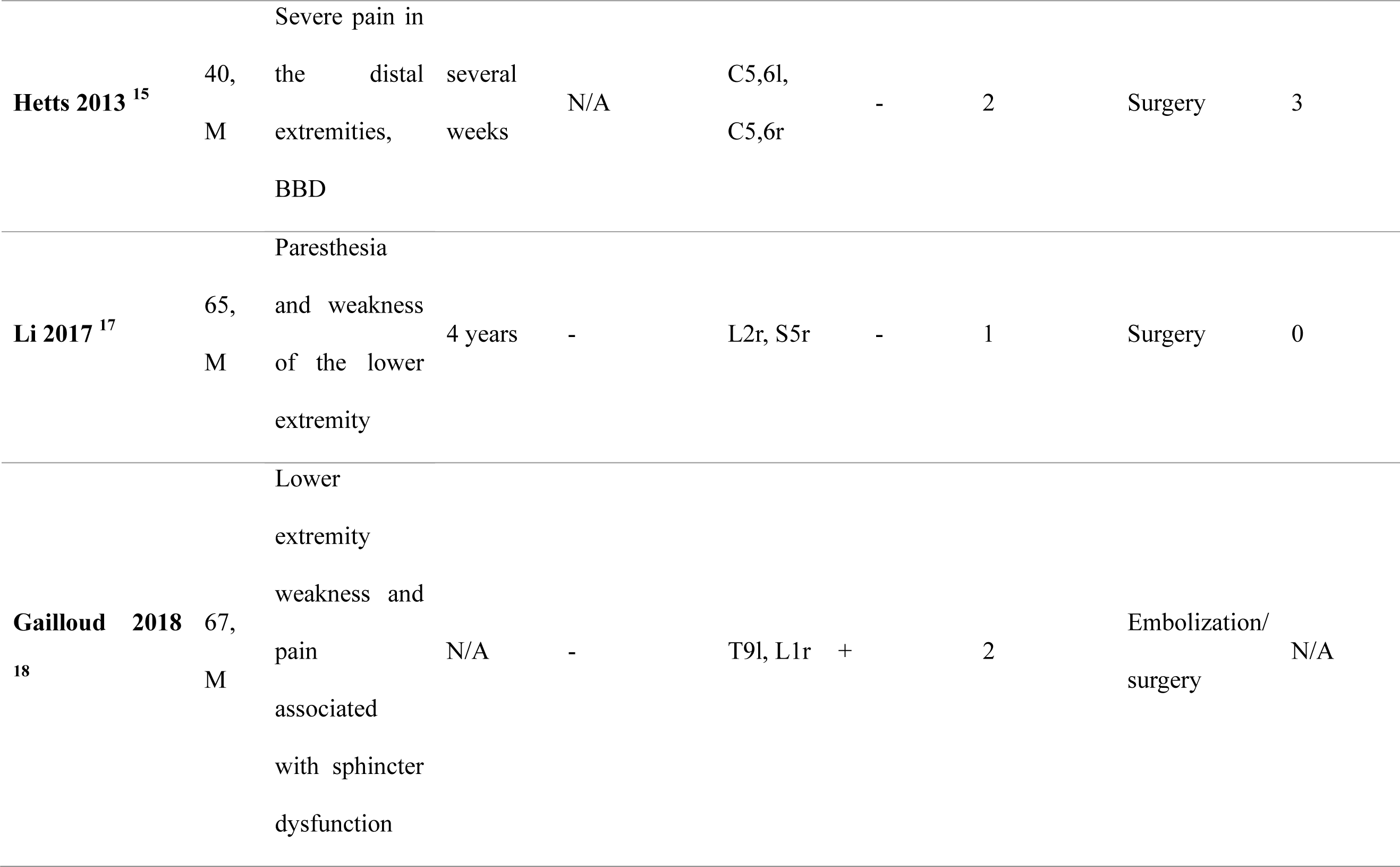
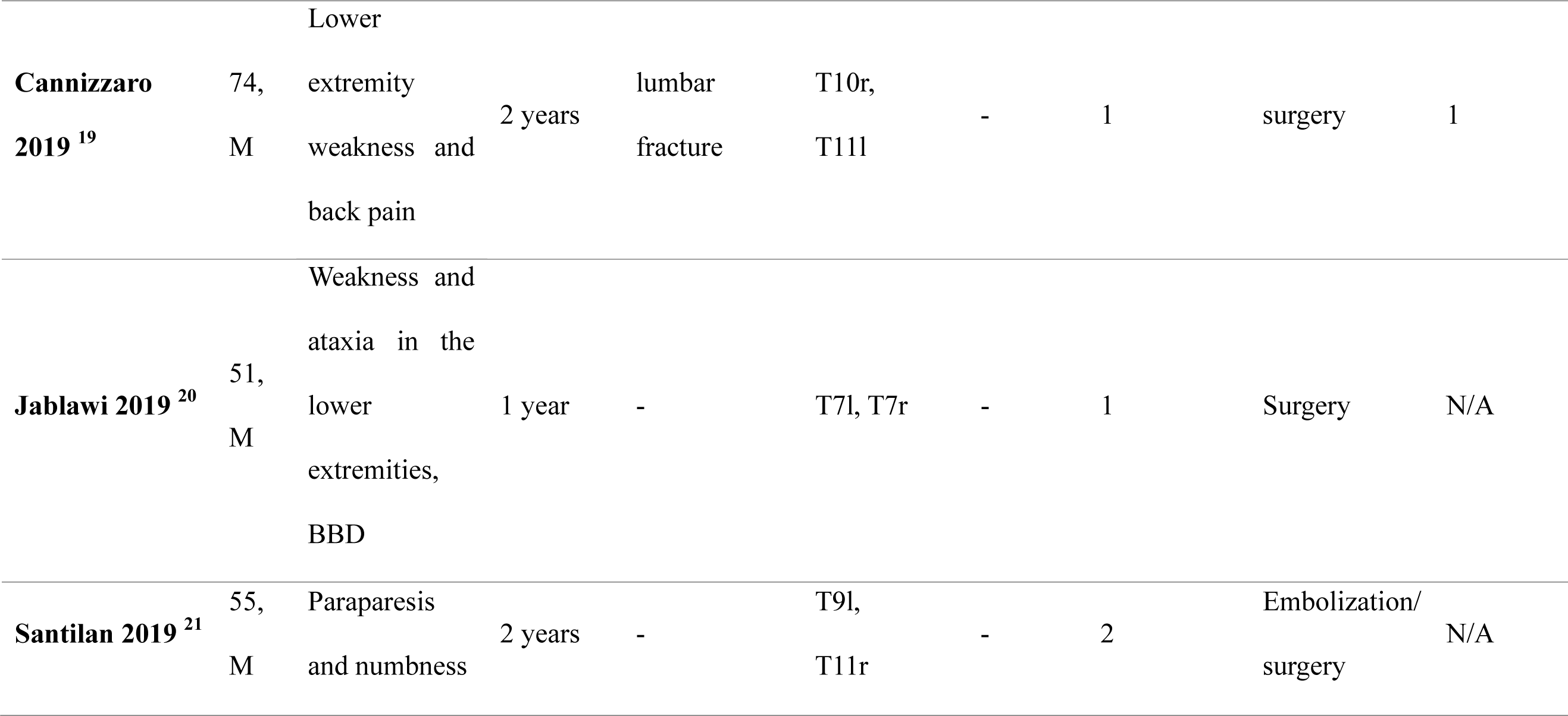
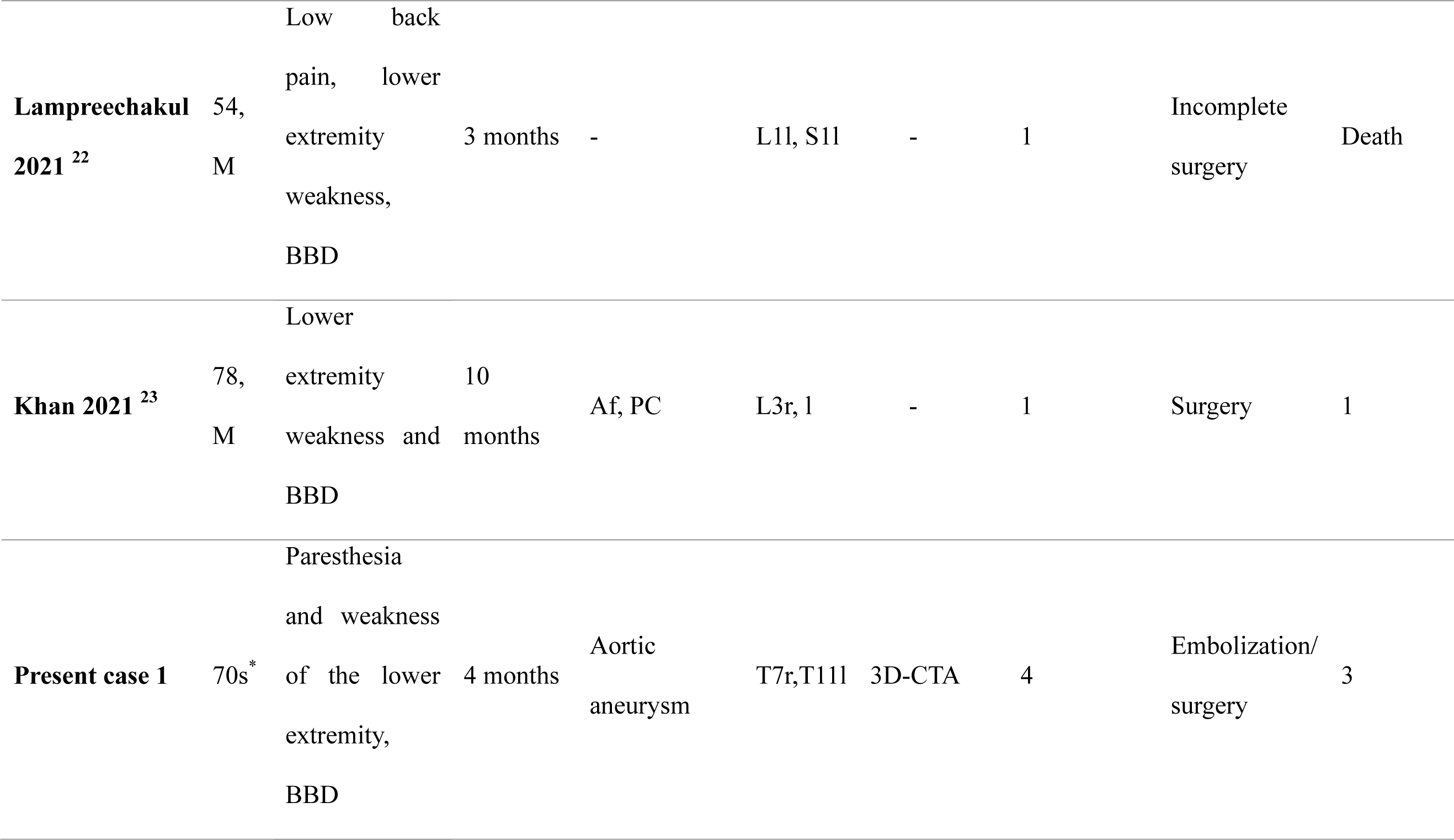
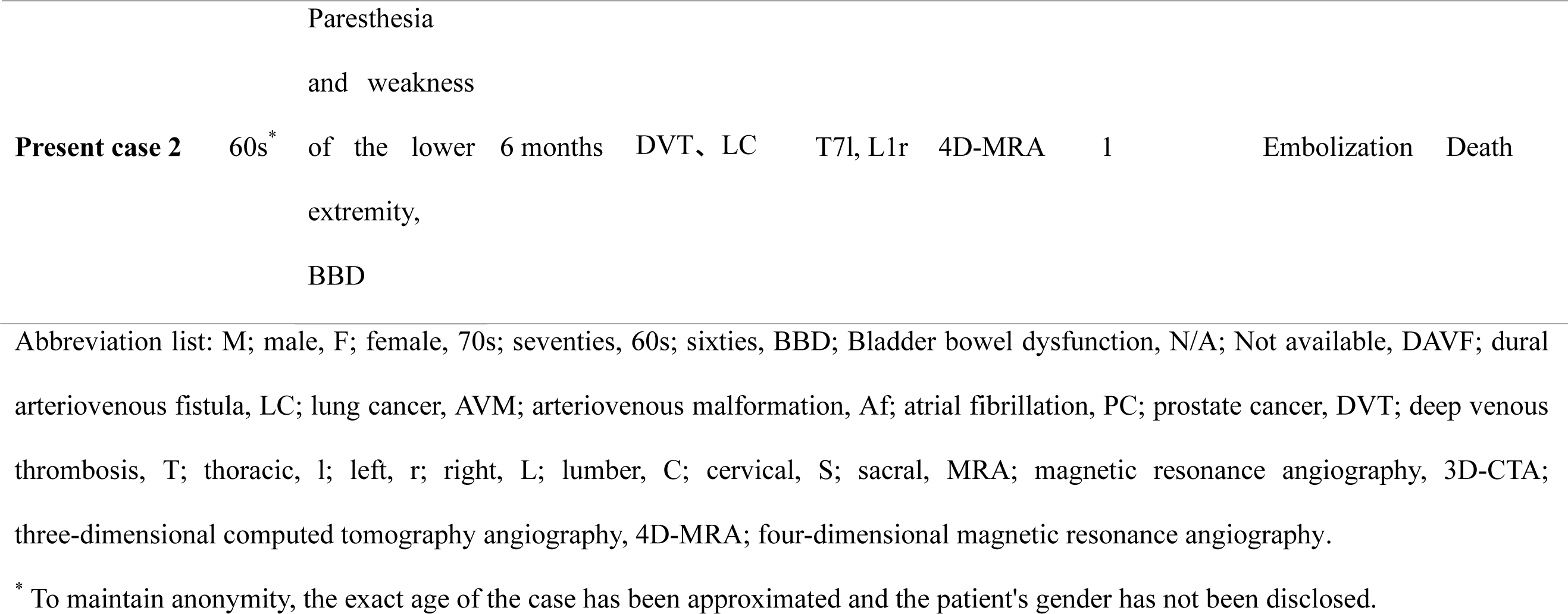
Literature review of 23 patients with multiple synchronous spinal arteriovenous fistulas.

### Patient population

Patients with SDAVF who were treated at Kobe University Hospital were prospectively registered in the clinical database. The database was reviewed to identify all patients with multiple SDAVFs who were treated by the senior author (F.A.) between 2008 and 2022. The patients were informed about available treatment options for these lesions, including conservative (nonoperative) management, endovascular embolization (if possible), and microsurgical resection. During the 14-year review period, the senior author operated on 18 patients with SDAVFs, 2 of whom had multiple SDAVFs.

### Statistical analyses

All statistical analyses were performed using the R software (version 3.5.3; R Foundation for Statistical Computing, Vienna, Austria).^23^ Data are presented as medians and standard deviations. The distribution of patients’ baseline characteristics was evaluated between the groups using descriptive statistics. The chi-square and Fisher’s exact tests were used for paired data to test for differences in distribution between the groups. Statistical significance was set at P values ≤0.05. All tests were two tailed.

### Ethical considerations

This study was approved by the Institutional Review Board of the Kobe University Hospital (approval number: #B230090). The study was conducted according to the principles of the Declaration of Helsinki regarding experimentation on human subjects and compliance with national regulations.

## RESULTS

### Demographics and presentation

A total of 23 male patients were included in this study (Figure 1). Their age ranged between 30 and 78 years, with a median age of 58.3 years. Among these patients, 18 (78.2%) presented with weakness of the lower extremities; 13 (56.5%) had paresthesia, sensory loss, or pain in the lower extremities; 10 (43.5%) presented with bladder and bowel dysfunction; and 1 (4.3%) was found incidentally. The duration from symptom onset to diagnosis was recorded in 21 of 23 cases. Fifteen (71.4%) of these 21 patients demonstrated symptoms for >6 months before the diagnosis. Regarding comorbidities, two (8.7%) patients had vascular malformations at other sites and three (13.0%) had malignant tumors.

### Neurological examination

SDAVFs were localized before spinal angiography in only five (21.7%) cases in our review, which included one case from the study by Dam-Hieu et al.,^8^ one from the study by Hanakita et al.[13], one from the study by Li et al.,^16^ one using magnetic resonance imaging (MRI) from the study by Khan et al.,^22^ and one (case #2) from the present study using four-dimensional (4D)-MRA. All SDAVFs were found using spinal angiography before embolization/surgery in 16 (69.6%) patients, whereas SDAVFs were newly found after embolization/surgery in 7 (30.4%).

The locations of all multiple SDAVFs are shown in Figure 2A. Twelve of the 23 (52.2%) patients had remote SDAVFs separated by more than three vertebral levels and constituted the remote SDAVF group. The remaining 11 patients were assigned to the non-remote SDAVF group (Figure 2B). All SDAVFs could not be identified at the time of treatment in five and two patients in the remote and non-remote SDAVF groups, respectively. SDAVFs in two patients in the non-remote SDAVF group at the time of treatment could not be identified because spinal angiography was only performed between the upper and lower vertebrae in one case and spinal angiography was not performed at the author’s hospital because of a referral from another institution in the other case. Among 12 patients with remote SDAVFs, SDAVFs were identified at the time of treatment in 7. SDAVFs were identified using whole-spinal angiography in two patients. Furthermore, three patients were investigated and were found to have multiple suspected shunts based on MRI and MRA results. Finally, 4D-MRA suggested the presence of SDAVFs in two patients, including one (case #2) in the present study.

**Figure 2.**
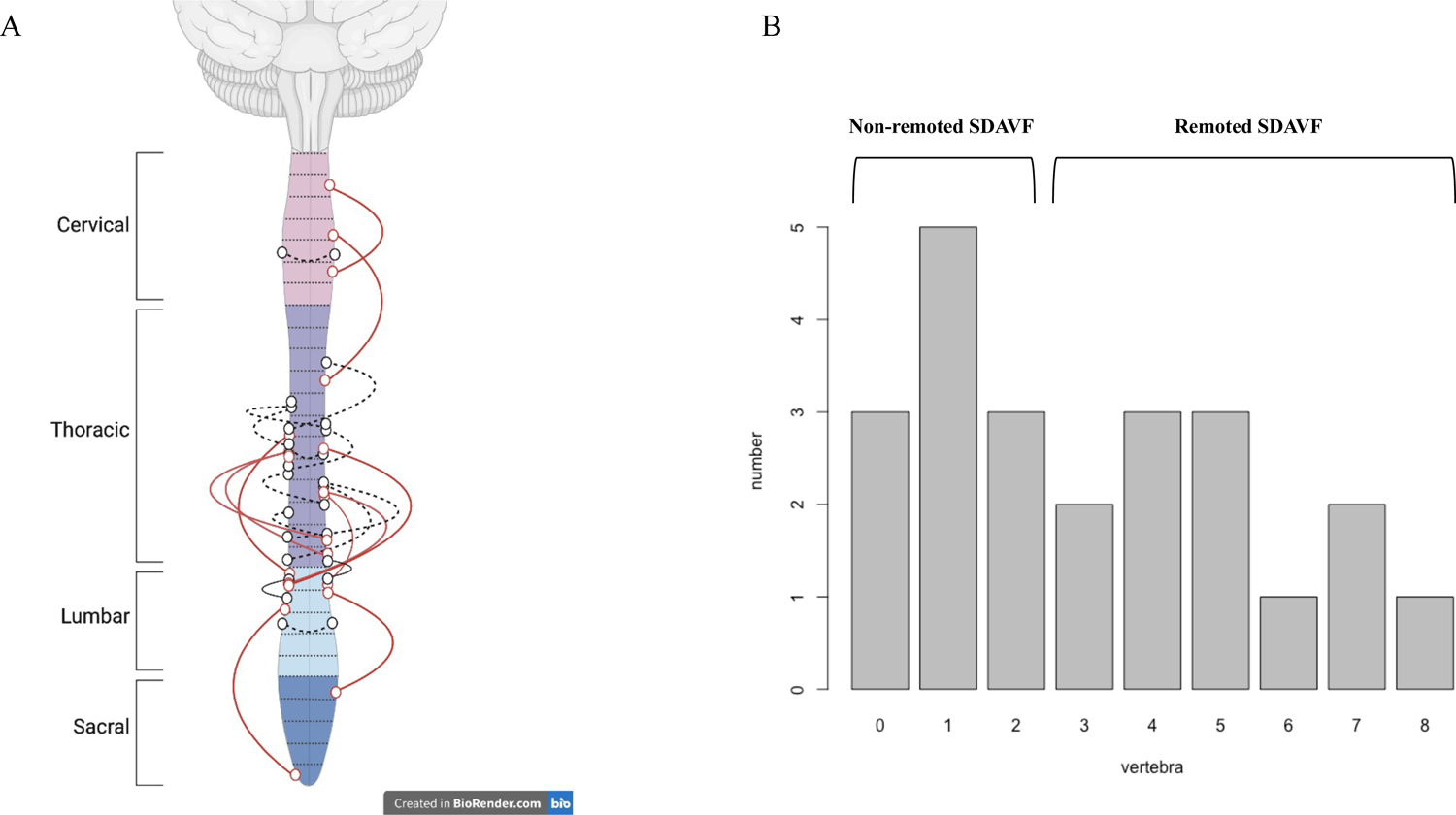
A: Locations of all SDAVFs. Dotted lines represent bilateral lesions, whereas solid lines represent ipsilateral lesions. The red line denotes lesions separated by more than three vertebral levels, whereas the black lines indicate lesions within a range of three vertebral levels. This figure was created with BioRender.com. B: Bar graph depicting the distribution of SDAVF cases based on the number of remote vertebral levels. Patients with lesions separated by more than three vertebral levels are defined to have remote SDAVFs.

### Surgical treatment and outcome

Among 23 patients, 11 underwent surgery only, 9 underwent surgery and received endovascular treatment, and 3 received endovascular treatment only. In the group with non-remote SDAVFs, the average number of treatments was 1.45. Conversely, in the group with remote SDAVFs, the average number of treatments was 1.91, with two cases resulting in incomplete treatment (Table 2).

**Table 2.** Univariate analysis between non-remote spinal dural arteriovenous fistulas (SDAVFs) and remoted SDAVFs.

|  | Non-remote<br>SDAVFs<br>(N=11) | Remote<br>SDAVFs<br>(N=12) | P value |
| --- | --- | --- | --- |
| Diagnosis failed rate – no. (%) | 2 (18.2) | 5 (41.7) | 0.44 |
| Treatment times – (ave) | 1.45 | 1.91 | 0.26 |
| Poor outcome – no. (%) | 1/7 | 8/11 | 0.049 |

Poor outcome: Modified Aminoff-Logue scale $\geq 2$ or death at 6 months
Abbreviation list: SDAVFs, spinal dural arteriovenous fistulas; ave, average

Among the 23 patients, the prognoses in 18 were reported in the articles. These prognoses were determined based on descriptions using a modified Aminoff-Logue scale and verified by two doctors. A modified Aminoff-Logue scale score was determined using information obtained 6 months post-treatment. When data from the 6-month post-treatment period were unavailable, the final outcome was determined based on prognosis at the final follow-up. A good prognosis was characterized by a score of 0–1 on the Aminoff-Logue scale, while poor prognosis was defined by a score of 2–5 or death at 6 months post-treatment. Among 23 patients, 8 of 9 with poor prognosis had remote SDAVFs. The remote SDAVF group tended to have significantly more cases of poor prognoses than the non-remote SDAVF group (1/7 vs. 8/11, P=0.049).

### Illustrative case (case #2 in the present study)

A patient in their sixties presented with paresthesia and motor deficits in the lower limbs. Their Aminoff-Logue gait scale classification was grade 4, and they reported an episode of urinary incontinence. Their medical history included deep vein thrombosis, stage IV lung cancer according to the tumor, node, metastasis classification, and bone metastasis. Cancer was classified as a stable disease after chemotherapy, and the patient’s life expectancy was estimated to be at least 6 months.

MRI revealed T2 hyperintensity in the mid-thoracic region (Th3-10) caudad and multiple dilated vessel flow voids in the thoracic region (Figure 3A). Owing to the patient’s allergy to iodinated contrast agents, 4D-CTA was not performed. Instead, 4D-MRA was used to localize the vascular shunts to the left T7 and right L1 regions (Figure 3B, C, D). All 4D-MRA examinations were performed using a 3-T MRI system (MAGNETOM Skyra; Siemens Healthcare, Erlangen, Germany) with a 32-channel head coil. The imaging parameters for 4D-MRA are listed in Table 3. Contrast injection was administered (0.2 mL per kg body weight dose of gadolinium-based contrast agents at a flow rate of 3 mL/s), followed by 20 mL of normal saline at the same rate. Selective digital subtraction angiography with bilateral injections from T6 to L2 revealed a right L1 SDAVF and a left T7 SDAVF (Figure 3E, G).

**Figure 3.**
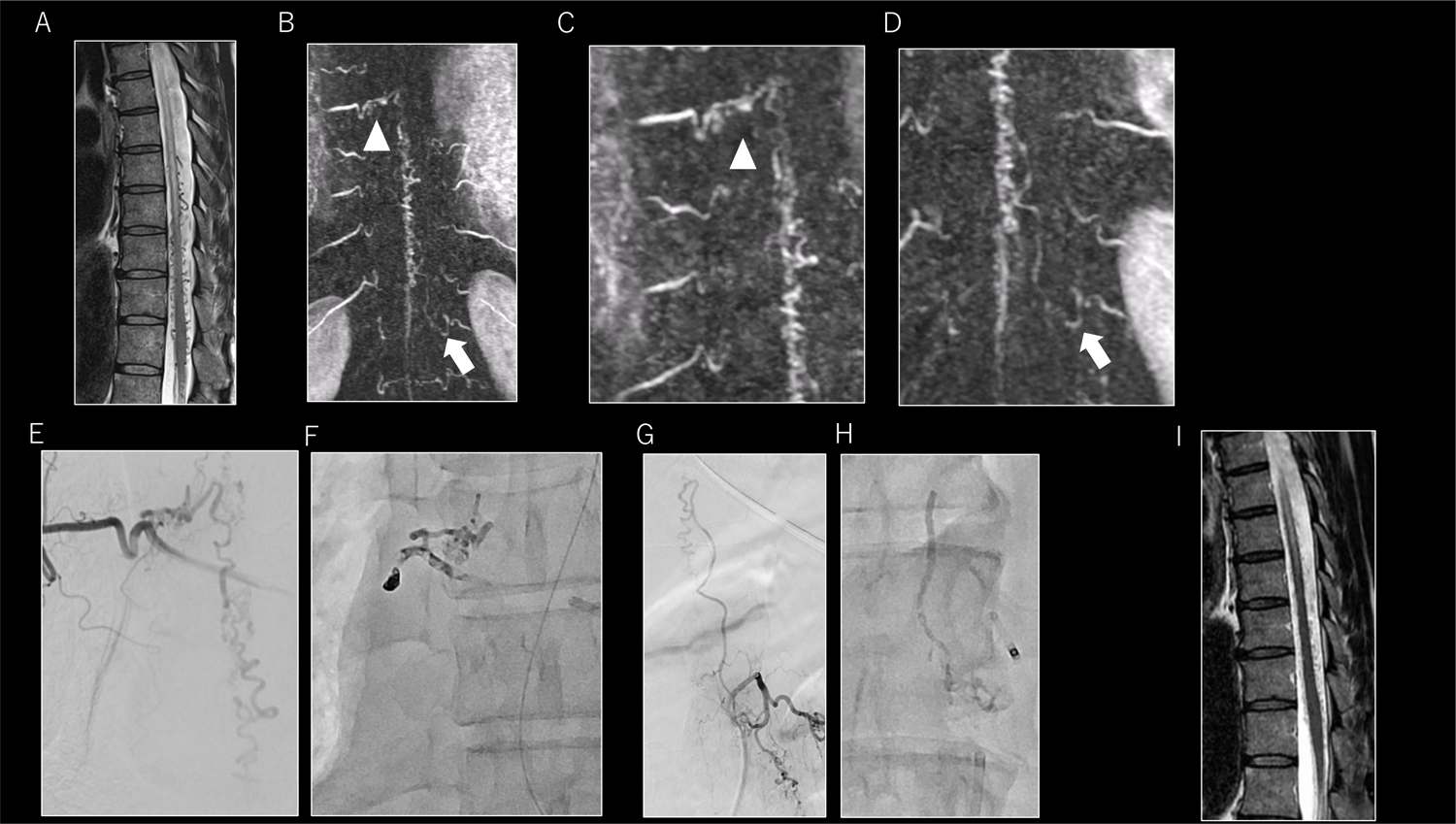
A: Magnetic resonance imaging (MRI) reveals T2 hyperintensity from the mid-thoracic region caudad and multiple dilated vessel flow voids in the thoracic region. B, C, D: 4D magnetic resonance angiography (4D-MRA) localizes vascular shunts to the left T7 and right L1 regions. B: General view of the lesion; C, D: Magnified images; C shows a T7 lesion, whereas D shows an L1 lesion. E, G: Selective DSA with bilateral injections from T6 to L2 reveals a left T7 SDAVF (E) and a right L1 SDAVF (G). F, H: The T7 and L1 feeding arteries are embolized using 12.5% NBCA, and subsequent spinal angiography demonstrates complete occlusion of the fistula. I: Follow-up MRI showing improved cord signal changes and resolution of dorsal dilated vessel flow voids.

**Table 3.**
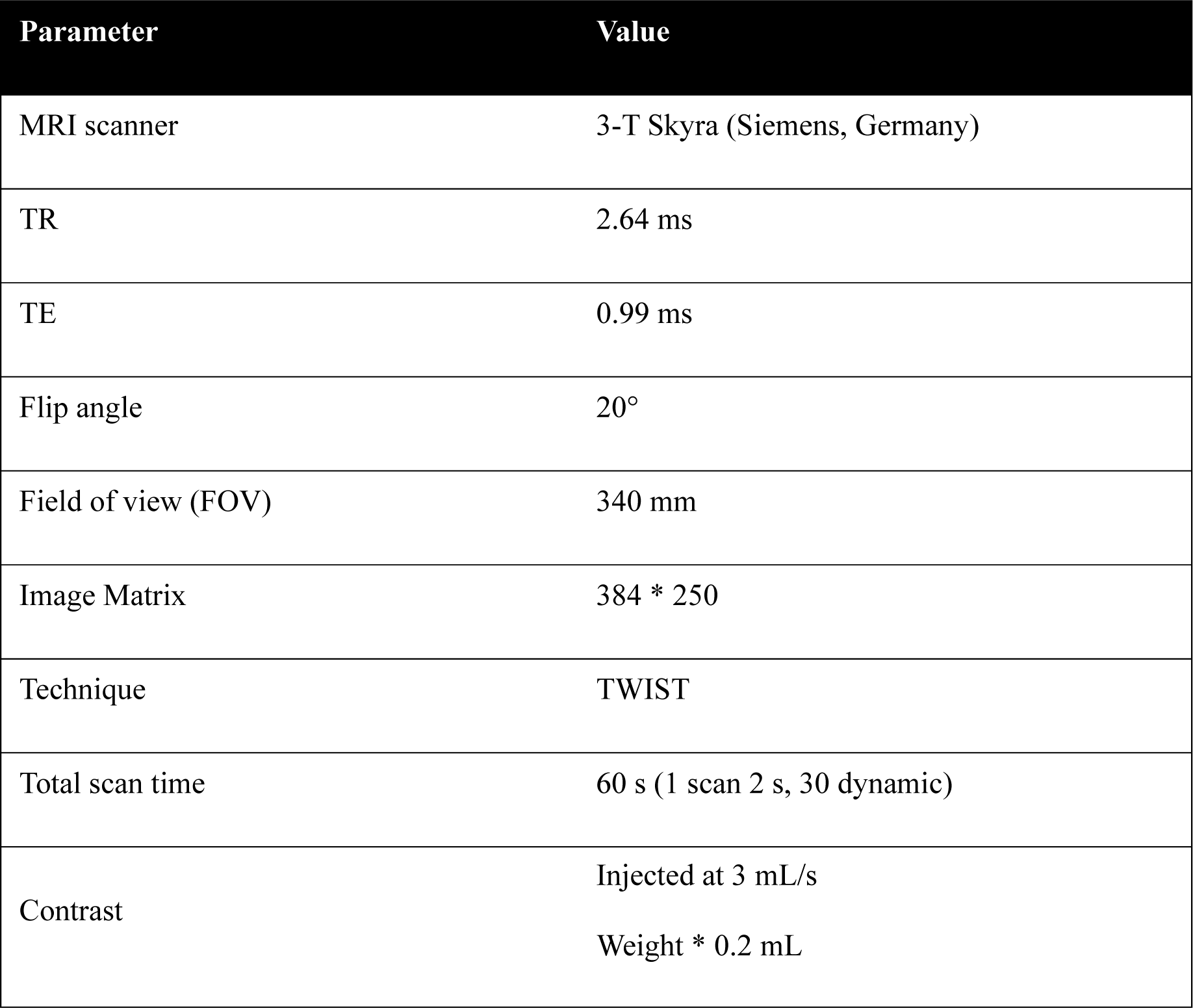
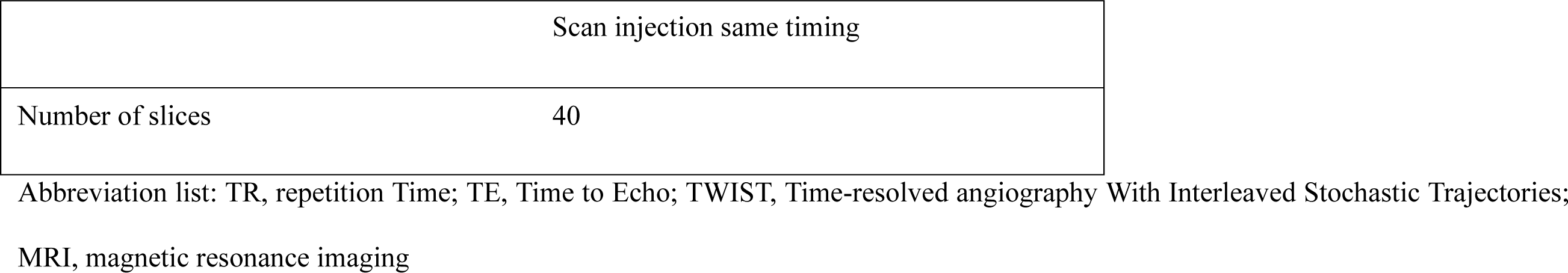
Imaging parameters for four-dimensional magnetic resonance angiography (4D-MRA)

The T7 and L1 feeding arteries were embolized using 12.5% n-butyl-2-cyanoacrylate, and subsequent spinal angiography demonstrated complete occlusion of the fistula (Figure 3F, H). Paresthesia was improved in both the lower legs; however, paralysis of the lower extremities worsened temporarily.

Follow-up MRI of the spine showed improved cord signal changes and resolution of the dorsal dilated vessel flow voids (Figure 3I). The patient died 3 months later because of lung cancer progression.

## DISCUSSION

To the best of our knowledge, this is the first systematic review of multiple synchronous SDAVFs. Multiple synchronous SDAVFs are rare; however, they have been reported previously. Prior studies suggested that multiple SDAVFs typically occur within a range of three or fewer vertebral levels.^24^ In contrast, more than 50% of SDAVFs identified in the present review were separated by more than three vertebral levels. Diagnosing multiple remote synchronous SDAVFs is challenging and often requires multiple treatment approaches. Localization of the fistula before spinal angiography is crucial for an accurate diagnosis. In case #2 in the present study, 4D-MRA was highly effective for localization. Considering the possibility of multiple remote SDAVFs, careful interpretation of imaging findings is essential for accurate diagnosis and appropriate treatment of these complex cases.

Multiple SDAVFs have seldom been reported in the literature. Pierot et al. identified 2 of 50 patients,^3^ van Dijk et al. found 1 of 49,^4^ and Krings et al. reported 1 of 129 patients having multiple SDAVFs.^25^ The estimated incidence of dual SDAVFs is quite low at approximately 1–2%. Therefore, in the present study, we focused solely on reports of synchronous SDAVFs and conducted a systematic review to address this issue.

All patients with multiple SDAVFs included in the previous report were male. In previous reports, SDAVF has been predominantly occurring in middle-aged men.^19^ The multiple occurrences of SDAVFs may be attributed to a systemic predisposition. Male sex hormones, such as androgens, are known to affect vascular function and structure and may play a role in the development of multiple SDAVFs.^26^

In the present review, the average duration from symptom onset to diagnosis was 15.6 months. Recently, a major tertiary hospital reported a mean symptom duration of 7 months.^27^ The development of multiple SDAVFs could be attributed to the extended duration from symptom onset to diagnosis.

Kaku et al. reported that most multiple SDAVFs exhibited similar outflow routes at neighboring levels.^24^ Notably, in the present review, >50% (12/23) of the SDAVFs were remote and separated by more than three vertebral levels. In the present review, patients with remote SDAVFs tended to receive more treatments and had significantly worse prognoses than those with non-remote SDAVFs. All cases of incomplete treatment were assigned to the remote SDAVF group. Since previous reports have indicated that longer time-to-treatment results in worse prognosis, the delay in improvement due to multiple treatments and the aggressiveness of the treatment may have contributed to the worsened prognosis.^28^

The gold standard for diagnosing SDAVF is spinal angiography, which confirms the diagnosis and reveals the following SDAVF characteristics before treatment: level of the fistula, anterior or posterior location to the spinal cord, venous drainage, and identification of normal spinal vasculature.^29^ During spinal angiography, neuroradiologists often cease their search after identifying a single fistula, potentially overlooking patients with multiple SDAVFs. However, localizing SDAVFs using catheter angiography may require approximately 40 selective injections into the intercostal, lumbar, sacral, cervical, and intracranial arteries.^30^ Whole-spinal angiography significantly burdens patients, with the percentage risk of permanent and transient neurological deficits being 0.2–1.0% and 4.6%, respectively.^31^ Jablawi et al. argued that whole-spinal angiography is not routinely warranted because of the extremely low incidence of multiple SDAVFs.^19^ A previous study reported that multiple SDAVFs often occur within a range of three or fewer vertebral levels.^24^ Therefore, further injections into the neighboring segmental arteries around the identified fistula zone are recommended upon detection of an SDAVF.

Moreover, pre-angiographic lesion localization using MRA or 3D-CTA has been reported to be beneficial for accurate diagnosis in SDAVF^32,33^; however, lesions may be missed in approximately 10% of cases.^19^ Accurate diagnosis of multiple SDAVFs may be particularly challenging because of the complex vascular conditions involved. Therefore, 4D-CTA has been reported to play a complementary role in diagnosing multiple SDAVFs.^32,33^ However, it has some limitations such as a constrained field of view (FOV) as narrow as 160 mm, which might not accurately visualize all remote SDAVFs that are located more than three vertebral levels away from the initially identified one. In the current case, multiple remote SDAVFs occurred synchronously and were separated by more than three vertebral levels, making diagnosis and treatment difficult. In recent years, 4D-MRA has been reported to be beneficial in identifying feeding arteries and accurately classifying intracranial dural AVFs. Furthermore, 4D-MRA has a broader FOV of approximately 340 mm compared with 4D-CTA, making it a valuable tool for diagnosing remote SDAVFs. The height of a single vertebral body is approximately 3 cm, which varies depending on the region. This implies that an imaging range of ≥15 cm is required to include both the upper and lower vertebral bodies when focusing on SDAVFs. Considering that the FOV of 4D-CTA is 16 cm, if the primary SDAVF is centered, the secondary lesion may fall outside the imaging range. Therefore, employing 4D-MRA with an FOV of 34 cm is considered highly advantageous in such cases. In case #2 in the present study, 4D-MRA could effectively localize remote SDAVFs. Yamaguchi et al. highlighted that the high radiation exposure dose is a significant limitation of 4D-CTA.^32,33^ Conversely, 4D-MRA poses no risk of radiation exposure, which is an advantageous feature of this imaging technique.

A single SDAVF can promote the development of a nearby second fistula owing to increased medullary venous pressure, concomitant venous stagnation, and thrombosis in the adjacent veins.^19^ However, in the present review, >50% of the SDAVFs were located at distant sites and separated by more than three intervertebral levels. Thus, the possibility of multiple remote SDAVFs should be considered when symptoms do not improve after treatment. Moreover, SDAVF may occur after thrombosis or in a hypercoagulable state.^34^ In the present review, three patients had deep venous thrombosis (DVT), atrial fibrillation, or neoplastic disease, predisposing them to thrombosis. The number of lower extremity paresis cases was high (18/23), suggesting that the number of unreported DVT cases might be higher. A systemic predisposition to thrombosis and prolonged duration from symptom onset to diagnosis may be associated with the development of multiple remote SDAVFs.

The present study is the first to focus on difficulties in treating and diagnosing remote synchronous multiple SDAVFs. Estimating the localization of lesions using MRA or CTA before spinal angiography helps diagnose and treat multiple remote SDAVFs. Furthermore, 4D-MRA may be particularly effective in diagnosing remote SDAVFs owing to its broad FOV and no risk of radiation exposure. Multiple synchronous SDAVFs are rare; however, more cases need to be evaluated to improve diagnostic and therapeutic approaches.

The present study had some limitations, which included a small number of case reports from a single institution and reliance on a literature review. Multiple SDAVFs are rare, and their detailed natural histories remain unknown. Moreover, some patients may have been excluded because of unconfirmed diagnoses or lack of additional treatment for multiple SDAVFs. There was also a potential for literature bias in this review. Finally, surgeons and neuroendovascular specialists should be aware that multiple SDAVFs can coincide in remote locations. Given the extreme rarity of multiple SDAVFs, it is believed that a multicenter, prospective registry study is essential.

In conclusion, multiple synchronous SDAVFs are rare. They were previously believed to be contiguous; however, >50% of the cases in this review involved remote lesions separated by more than three intervertebral levels. Diagnosing multiple remote SDAVFs is challenging, and multiple treatments are often necessary. Consequently, localizing the fistulas before spinal angiography is crucial in managing multiple remote SDAVFs, and 4D-MRA proved beneficial in our case. Considering the possibility of multiple remote SDAVFs, careful interpretation of imaging findings is essential for accurate diagnosis and appropriate treatment planning.

## List of abbreviations

CTA: Computed tomography angiography
DVT: Deep venous thrombosis
FOV: Field of view
MRA: Magnetic resonance angiography
MRI: Magnetic resonance imaging
PRISMA: Preferred Reporting Items for Systematic Reviews and Meta-Analyses
SDAVF: Spinal dural arteriovenous fistula
4D: Four dimensional

## Acknowledgments

None.

## Data availability statement

This review was not registered and the review protocol was not prepared. All data generated or analyzed during this study are included in this article.

